# Effectiveness of Smoking Cessation Interventions in People With Cancer: A Systematic Review and Meta-analysis of Randomized Controlled Trials

**DOI:** 10.1101/2025.01.11.25320383

**Authors:** Livingstone Aduse-Poku, Hui G. Cheng, Oxana Palesh, Susan Hong

## Abstract

**Introduction:** Smoking at or following a cancer diagnosis increases both all-cause and cancer-specific mortality, adverse treatment outcomes, as well as the risk for disease progression and tobacco-related second primary cancers. This protocol outlines a systematic review and meta-analysis evaluating the effectiveness of smoking cessation interventions in cancer patients based on randomized controlled trials (RCTs), delivering the highest quality of evidence for causal inference.

**Methods:** A systematic search of the literature will be conducted across PsychInfo, EMBASE (through OVID), PubMed (also through OVID), and the Cumulative Index to Nursing and Allied Health Literature (CINAHL). RCTs published in English, designed to assist adult cancer survivors who smoke to quit, regardless of their cancer type, stage, or treatment, will be included. Title and abstract screening will be carried out independently by two authors. All potentially eligible full-text articles will be independently reviewed. Any disagreements will be resolved through discussion. Extracted data will encompass study characteristics, intervention details, participant numbers and attributes, outcomes, quit attempts, and follow-up duration. We will evaluate the risk of bias using the Cochrane Risk of Bias Tool 2 (ROB-2). To evaluate the outcomes of the combined studies, a meta-analysis and/or network meta-analysis will be performed with both fixed-effect models and random-effects models using R.

**Conclusion:** This systematic review will address a critical research gap and provide tailored insights for smoking cessation among cancer survivors.

## Introduction

Smoking is a highly prevalent and preventable risk factor for many diseases and premature death.^1^ Globally, there are more than 1.1 billion smokers, and more than seven million smokers are killed by smoking each year.^2^ The leading causes of mortality associated with tobacco smoking include cancer, coronary heart disease, stroke, and chronic obstructive pulmonary disease.^3^ According to the United States National Institute of Health (NIH), smoking is the leading cause of cancer and cancer-related deaths, accounting for nearly 10 million deaths yearly.^4^

Smoking at or following a cancer diagnosis increases both all-cause and cancer-specific mortality, adverse treatment outcomes, as well as the risk for disease progression and tobacco-related second primary cancers.^5,6^ The overall mortality rate is 4% and 20% higher for patients who continue to smoke after cancer diagnosis compared to those who had quit.^7,8^ Consequently, offering cessation assistance to cancer patients who smoke is important for improving their prognosis and quality of life. Despite the numerous health complications that can arise from smoking, 33% to 50% of cancer patients continue to smoke or relapse after attempting to quit.^9,10^ Cancer patients experience heightened stressors, such as depression and anxiety, which significantly hinder their efforts to quit smoking and increase the risk of relapse^11,12^ A poorer prognosis does not necessarily motivate patients to quit smoking; evidence suggests that individuals are more likely to discontinue the habit if the cancer is causally linked to their smoking behavior.^13,14^ A significant proportion of patients who relapse into smoking after surgical treatment for early-stage non-small-cell lung cancer do so within the first 1–6 months post-surgery.^15^

Quitting smoking is the most efficacious means to mitigate smoking-related premature mortality and morbidity. Smoking cessation interventions can be classified into behavioral and pharmacological approaches. Behavioral and pharmacological treatments are postulated to possess synergistic mechanisms of action and to enhance the likelihood of sustaining long-term abstinence independently. Pharmacological interventions involve the use of medications like nicotine replacement therapy, varenicline, and bupropion. These medications have been shown to increase the risk of quitting smoking.^11,16^ Behavioral interventions encompass cognitive-behavioral therapy, motivational interviewing, telephone counseling, individual or group counseling, and web-based interventions.^17^ Other interventions involve the combination of both pharmacological and behavioral interventions.^18^

Extensive research has been conducted to determine the most effective interventions for helping healthy individuals quit smoking. A considerable body of systematic reviews, comprising findings from more than 600 studies, have been conducted to elucidate the most effective smoking cessation approaches for this population.^19–24^ Yet, the relevance of these insights to cancer survivors remains uncertain. A limited number of studies have specifically evaluated the effectiveness of these programs among oncology population. Many of the existing studies on smoking cessation programs for cancer patients have utilized observational designs to assess outcomes. Two systematic reviews that have examined effective smoking cessation programs among cancer patients included observational studies, and therefore, could not draw strong conclusions on the causal role and effectiveness of smoking cessation interventions.^25,26^

A recent systematic review, which included both observational studies and Randomized Controlled Trials (RCTs), found that a combination of pharmacological and behavioral interventions may be the most efficacious intervention for smoking cessation among cancer survivors.^25^ This systematic review and meta-analysis is of particular relevance because it will be the first to evaluate the effectiveness of smoking cessation interventions in cancer patients based on experimental studies (e.g., RCTs), delivering the highest quality of evidence for causal inference. This systematic review will address a critical research gap and provide tailored insights for smoking cessation among cancer survivors. Our results will inform clinical practice, policymaking, and the future research needed to optimize care for cancer survivors, leading to advances in oncology care and public health.

## Methods

This systematic review will be performed in accordance with a predetermined protocol and will be reported to be consistent with the Preferred Reporting Items for Systematic Reviews and Meta-Analysis (PRISMA) statement.^27^

### Search strategy

The search strategy will include the use of search terms, both primary keywords and specific vocabulary terms (see Appendix) for controlled searches MESH and EMTREE, across four databases: PsychInfo, EMBASE (through OVID), PubMed (also through OVID), and the Cumulative Index to Nursing and Allied Health Literature (CINAHL).

### Eligibility criteria

We plan to include studies that examined interventions designed to assist adult cancer survivors who smoke to quit, regardless of their cancer type, stage, or treatment. We will only consider randomized controlled trials, including cluster randomized controlled trials. Comparison groups for the included trials may comprise no intervention controls, usual practice, or alternative interventions. Quasi-experimental trials with comparison/control groups, including non-randomized pre-post trials with one or more intervention and control groups, time-series/interrupted time-series trials with independent control groups, preference trials, and regression discontinuity trials, will be excluded. Research articles published up to October 2024 will be included, with a preference for full-length articles in English from peer-reviewed sources.

### Participants

The included studies will involve adult participants diagnosed with any type of cancer, excluding non-melanoma skin cancer, who are current smokers. There will be no limitations on the type or stage of treatment. Studies investigating a diverse population of cancer patients will be eligible for inclusion.

### Types of interventions

Interventions that aim to improve the smoking cessation outcomes of patients with cancer will be included. Interventions will be categorized into three main groups: behavioral approaches (such as cognitive therapy and motivational interviewing), medication-based approaches (like bupropion, varenicline, nicotine replacement therapy, or a mix), and combined approaches (which involve any behavioral approach with a medication approach).

### Outcome

The primary outcome of interest in these studies will be measures of smoking cessation. Trials must provide a quantifiable measurement of abstinence from smoking to be included. This could encompass point prevalence rates, continuous abstinence, or current smoking status. Given the lack of a safe level of tobacco consumption, smoking reduction will not be considered a valid outcome. Studies will only be included if they clearly define smoking cessation, accepting both self-reported abstinence and biochemical verification, such as carbon monoxide levels in exhaled air or blood nicotine concentrations.

### Selection of Studies and Data Extraction

Database searches will be conducted by LAP. After deduplicating references, they will be loaded into the Covidence platform. Title and abstract screening will be carried out independently by two authors, LAP and HC. All potentially eligible full-text articles will be independently reviewed by LAP and HC. Any disagreements will be resolved through discussion. Extracted data will encompass study characteristics, intervention details, participant number and attributes, outcomes, quit attempts, and follow-up duration. Two authors, LAP and HC, will independently extract data from the included studies.

### Assessment of risk of bias in included studies

We will evaluate the risk of bias using the Cochrane Risk of Bias Tool 2 (ROB-2). We will assess each included study for the risk of bias across five domains: i. random sequence generation, ii. concealment of allocation (for cluster-randomized controlled trials, where participants were recruited after allocation to intervention or control status, we will consider whether individuals may have been selectively recruited or differentially refused to participate based on the known allocation, as this could introduce ascertainment bias), iii. performance bias iv. detection bias, and v. Attrition. Additionally, we will document any other potential sources of bias that do not fit within the above categories.

### Evidence synthesis

We will apply the GRADE methodology to assess the certainty of the evidence presented in the Cochrane reviews, incorporating the guidance for overviews provided by Cochrane in 2019 and utilizing the GRADEpro software. The certainty of the evidence will be downgraded or upgraded based on several factors. Factors that may lead to downgrading include risk of bias, inconsistency, imprecision, and publication bias. Conversely, factors that may result in upgrading the certainty include large effect size, dose-response gradient, and plausible confounding.

### Data analysis

To evaluate the outcome of the combined studies, a meta-analysis and/or network meta-analysis will be performed with both fixed effect models and random effect models using R. The inverse variance method will be used to calculate the risk ratio. We will perform sensitivity analysis based on intervention type monotherapeutic interventions (pharmacologic or behavioral interventions) and combined interventions (pharmacologic and behavioral interventions). Additionally, we will perform a sensitivity analysis based on a smoking cessation assessment (self-report only or with biochemical verification). A funnel plot will be generated to evaluate potential publication bias. The funnel plot will depict the standard error of the logarithmic risk ratio on the vertical axis and the risk ratio on the horizontal axis. Asymmetry will be assessed both visually and through the statistical Egger test, which will be performed in R Studio.

## Data Availability

This is a systematic review protocol

## SEARCH TERMS

For Medline and Embase

|  |  | Search terms |
| --- | --- | --- |
| Patients diagnosed with cancer | 1 | neoplasm.mp. or exp Neoplasms/ neoplasm\$.mp. |
|  | 2 | exp Breast Neoplasms/ or cancer patients.mp. |
| | 3 | cancer patient\$.mp. |
| | 4 | cancer survivor\$.mp. |
|  | 5 | cancer survivors.mp. or exp Cancer Survivors/ |
|  | 6 | exp Lung Neoplasms/ or lung cancer patients.mp. |
|  | 7 | exp Colorectal Neoplasms/ or colorectal cancer survivors.mp. |
|  | 8 | exp Prostatic Neoplasms/ or prostate cancer patients.mp. |
|  | 9 | 1 or 2 or 3 or 4 or 5 or 6 or 7 or 8 |
| Smoking cessation interventions / program / methods | 10 | Smoking Cessation.mp. or Smoking/ or exp Smoking Cessation/ or exp Smoking Prevention/ |
|  | 11 | Smoking Cessation programs.mp. |
| | 12 | Smoking Cessation program\$.mp. |
| | 13 | Smoking Cessation Agent\$.mp. |
|  | 14 | Nicotine Replacement Therapy.mp. |
|  | 15 | Bupropion.mp. |
|  | 16 | Varenicline.mp. or exp Varenicline/ |
|  | 17 | Behavior Therapy.mp. or exp Behavior Therapy/ |
| | 18 | Motivational Interview\$.mp. |
| | 19 | Self-Help Group\$.mp. |
| | 20 | Cognitive Behavioral Therap\$.mp. |
|  | 21 | exp Hypnosis/ or hypnosis.mp. |
| | 22 | comprehensive counsel\$.mp. |
|  | 23 | 10 or 11 or 12 or 13 or 14 or 15 or 16 or 17 or 18 or 19 or 20 or 21 or 22 |
| Randomized controlled trials/ interventional studies | 24 | Randomized Controlled Trial.mp. or exp Randomized Controlled Trial/ or exp Double-Blind Method/ or Doubleblind.mp. or Single-Blind Method/ or Single-blind.mp. or Random Allocation.mp. or Random Allocation/ or Clinical Trials.mp. or exp Clinical Trial/ |
| Combined | 34 | 9 AND 23 AND 24 |

For PubMed / CINAHL

#1 “Neoplasms”[Mesh Terms] OR neoplasm*[tiab] OR cancer*[tiab] OR malginan*[tiab] OR oncology[tiab] OR tumor[tiab] OR tumour[tiab] OR carcinoma[tiab]

#2 “Tobacco Smoking”[Mesh] OR “Cigarette Smoking”[Mesh] OR “Cigar Smoking”[Mesh] OR “Smoking”[Mesh] OR tobacco[tiab] OR Smoke*[tiab] OR cigarette*[tiab] OR cigar[tiab] OR smoking[tiab] OR “nicotine dependence”[tiab]

#3 “Smoking Cessation”[Mesh] OR “Tobacco Use Cessation Devices”[Mesh] OR “Smoking Cessation Agents” [Pharmacological Action] OR “Smoking Cessation Agents”[Mesh] OR “Tobacco Use Cessation”[Mesh] OR quit*[tiab] OR stop*[tiab] OR Cessation[tiab] OR “Smoking Cessation”[tiab] OR “Tobacco Use Cessation Device*”[tiab] OR “Smoking Cessation Agent*”[tiab] OR “Tobacco Use Cessation”[tiab] OR “brief physician advice”[tiab] OR “physician advice”[tiab] OR “smoking cessation advice”[tiab] OR counselling[tiab] OR “cognitive behavioral therapy”[tiab] OR “nurse intervention”[tiab] OR “motivational interviewing”[tiab] OR “Stage-based intervention*”[tiab] OR “Print-based self-help”[tiab] OR “group therapy”[tiab] OR exercise*[tiab] OR “financial incentive”[tiab] OR “monetary incentive”[tiab] OR hypnotherapy[tiab] OR “nicotine replacement therapy”[tiab] OR bupropion[tiab] OR nortriptyline[tiab] OR varenicline[tiab] OR cystine[tiab] OR e-cigarette[tiab] OR antidepressant*[tiab] OR “tricyclic antidepressant*”[tiab] OR “monoamine oxidase inhibitor*”[tiab] OR “selective serotonin reuptake inhibitor*”[tiab] OR “atypical antidepressant*”[tiab] OR “St. John’s wort”[tiab] OR anxiolytic*[tiab] OR buspirone[tiab] OR diazepam[tiab] OR doxepin[tiab] OR meprobamate[tiab] OR ondansetron[tiab] OR metoprolol[tiab] OR oxprenolol[tiab] OR propanolol[tiab] OR rimonabant[tiab] OR taranabant[tiab] OR clonidine[tiab] OR lobeline[tiab] OR dianicline[tiab] OR mecamylamine[tiab] OR nicobrevin[tiab] OR “nicotine vaccine*”[tiab] OR naltrexone[tiab] OR naloxone[tiab] OR buprenorphine[tiab] OR “silver acetate”[tiab]

#4 #1 and #2 and #3

